# Prediagnostic breast milk DNA methylation alterations in women who develop breast cancer

**DOI:** 10.1101/19001925

**Authors:** Lucas A Salas, Sara N. Lundgren, Eva P. Browne, Elizabeth C. Punska, Douglas L. Anderton, Margaret R Karagas, Kathleen F. Arcaro, Brock C. Christensen

## Abstract

**Background:** Prior candidate gene studies have shown tumor suppressor DNA methylation in breast milk related with history of breast biopsy, an established risk factor for breast cancer. To further establish the utility of breast milk as a tissue-specific biospecimen for investigations of breast carcinogenesis we measured genome-wide DNA methylation in breast milk from women with and without a diagnosis of breast cancer in two independent cohorts.

**Methods:** DNA methylation was assessed using Illumina HumanMethylation450k in 87 breast milk samples. After quality control, 368,171 autosomal CpG loci were analyzed. Cell type proportion estimates from RefFreeCellMix were calculated and adjusted for in this Epigenome Wide Association Study using linear mixed effects models adjusted for history of breast biopsy, age, time of delivery, cell type proportion estimates, array chip, and subject as random effect.

**Results:** Epigenome-wide analyses identified 58 differentially methylated CpG sites associated with a breast cancer diagnosis in the prospectively collected milk samples from the breast that would develop cancer compared with women without a diagnosis of breast cancer (*q*-value < 0.05). Nearly all CpG sites associated with a breast cancer diagnosis were hypomethylated in cases compared with controls, and were enriched for CpG islands. In addition, inferred repeat element methylation was lower in breast milk DNA from cases compared to controls, and cases exhibited increased estimated epigenetic mitotic tick rate as well as DNA methylation age compared with controls.

**Conclusion:** Breast milk has utility as a biospecimen for prospective assessment of disease risk, for understanding the underlying molecular basis of breast cancer risk factors, and improving primary and secondary prevention of breast cancer.

## BACKGROUND

Breast cancer is the most common non-keratinocyte cancer in women in the USA, with over 270,000 new cases each year [1]. Established risk factors for breast cancer include age, reproductive history, and family history of disease and can be used to estimate disease risk [2,3]. Additionally, and beyond the recognized role of inherited *BRCA* mutation, individual germline genetic variants, and even polygenic risk scores from genome-wide association studies have also contributed to breast cancer risk assessment [4–6]. Nonetheless, a large gap in the capacity to predict breast cancer risk remains, and the molecular basis of breast cancer risk and carcinogenesis has largely not been studied using target-organ biospecimens from premenopausal women.

Epigenome-wide association studies (EWAS), using surrogate tissues such as peripheral blood DNA, have also had some success testing the relation of DNA methylation with cancer risk [7–9]. However, unlike genetic variation and germline alterations that confer cancer risk, cytosine modifications that contribute to cancer risk as disease initiating and promoting events are overwhelmingly tissue specific. Defining and leveraging knowledge of tissue-specific early DNA methylation alterations for screening or risk models in normal, nontumor human tissues is challenging for most common tumor types. Yet, use of breast-specific substrate to investigate breast cancer risk has shown promise in early studies measuring cell composition, cytology, and candidate gene DNA methylation from nipple aspirate fluid, though as a substrate, nipple aspirate fluid can be challenging to obtain and typically yields very low volume [10–14]. Recently, the utility of altered DNA methylation in cancer screening and risk assessment was established in colon cancer as part of the Cologuard multi-target assay where a tissue-specific biospecimen (stool) is obtained and measured without using an invasive procedure [15].

The majority of extensive DNA methylation alterations observed in invasive breast cancer compared with normal breast tissue, are already present in pre-invasive disease [16], [17]. In addition, age-related variation in normal breast tissue DNA methylation has been shown to occur at CpG sites that are more likely to be altered in breast tumors [18], suggesting that early measures of DNA methylation in the pathologically normal breast has value as a biomarker for future breast cancer risk [18]. Typically, mammary epithelial cells cannot be accessed without invasive procedures (breast biopsy), lavage, or other relatively impractical methods. However, exfoliated mammary epithelial cells are abundant in breast milk [19], a tissue-specific substrate obtained without invasive procedure. These cells are an excellent target for biomarker development, and prior candidate gene studies have shown that methylation-induced silencing of tumor suppressor genes in breast milk is related with history of breast biopsy, an established risk factor for breast cancer [20–22]. Given that 85% of 40 year-old women in the USA have given birth [23], breast milk is a viable noninvasive source of mammary epithelial cells [24]. We investigate the relation of early epigenetic alterations with breast cancer risk using cells obtained from breast milk in controls compared with prospectively collected milk specimens from subjects who were later diagnosed with breast cancer.

## METHODS

### Study population

Two different study populations were included in this study: 1) women from the “Molecular Biomarkers for Assessing Breast-Cancer Risk” project at the University of Massachusetts Amherst (UMass), and 2) participants of the New Hampshire Birth Cohort study (NHBCS) at Dartmouth College. UMass subjects were women older than 18 years of age. They were either lactating or have recently given birth, and they had a history of either breast biopsy or breast cancer. UMass subjects were asked to provide one or two breast milk samples expressed in a single pumping session. NHBCS participants characteristics has been described previously [25]. Briefly, NHBCS eligibility criteria included: English speaking, literate, and mentally competent women carrying a singleton pregnancy, 18–45 years of age, and whose primary source of residential water was a private well. Women who planned to move during pregnancy were excluded from this study. NHBCS participants were asked to bring bilateral breast milk samples to the postpartum follow-up appointment. All study participants provided written informed consent prior to the study according to the guidelines of Institutional Review Board of the University of Massachusetts Amherst and the Committee for the Protection of Human Subjects at Dartmouth. Women in both studies were asked to complete a questionnaire about general health, reproductive health, and personal breast biopsy and breast cancer history. Each woman’s samples were classified into five different groups: (1) no breast cancer history, (2) healthy breast, contralateral breast cancer before donation, (3) ipsilateral breast cancer diagnosis before donation, (4) healthy breast, contralateral cancer diagnosis after donation, and (5) sample from the ipsilateral breast with cancer after donation. For this analysis, we report the results of model milk samples from control subjects and from subjects with a subsequent diagnosis of breast cancer.

### Sample collection

Using a previously described method [24], breast milk was processed within 24 hours of sample collection to obtain DNA. Briefly, DNA was extracted from 1 - 10 mL of milk from each breast and stored at −20 °C until DNA extraction.

### DNA extraction and genome-wide DNA methylation array

DNA was isolated using the Qiagen DNeasy Blood and Tissue Kit (Qiagen, Valencia, CA) and bisulfite converted using the EZ DNA Methylation kit (Zymo, Irvine, CA). Samples were randomized across several plates and subsequently subjected to epigenome-wide DNA methylation assessment using Illumina Infinium HumanMethylation450 BeadChip, which measured ∼485,000 CpG sites genome-wide (Illumina, San Diego, CA). Microarrays were processed at USC core facility following standard protocols. The data were assembled using GenomeStudio methylation software (Illumina, San Diego, CA) without normalization per the manufacturer’s instructions. The methylation status for each individual CpG locus (β-value) was calculated as the ratio of fluorescent signals (β = Max(M,0)/ [Max(M,0) + Max(U,0) + 100]), ranging from 0 (no methylation) to 1 (complete methylation) using the average probe intensity for the methylated (M) and unmethylated (U) alleles. We read the idat files using the minfi R package [26]. β-values were background corrected using methylumi-noob and normalized using functional normalization.[27] Our pipeline included array control probes to assess sample quality and evaluate potential problems such as poor bisulfite conversion or color-specific issues for each array as described previously [28,29]. All CpG loci on X and Y chromosomes, CpH, and loci with potential problems of cross-reactivity, tracking to polymorphisms with minor allele frequencies over 5% for the general population, or common copy number alterations,[30] were excluded from the analysis, leaving 368,171 autosomal CpG loci in 92 samples. Principal components analysis and multiple dimension scaling were used to identify potential technical batches. Additionally, we used a principal component regression analysis to investigate the top eight principal components in relation to potential batch-associated differences. Subjects with missing covariate data were excluded from modeling, resulting in 87 samples. DNA methylation β-values were logit_2_ transformed to M-values for the analyses [31].

### Cell mixture analysis

In order to identify and adjust for potential cell type heterogeneity in the breast milk samples we used a reference-free decomposition (RefFreeCellMix) of the DNA methylation matrix into cell-type distributions and cell-type methylomes, using the expression Y = ?*Ω^?^ [32]. We explored a range of k cell types from 2 to 10. Note that the decomposition will be based on Y, but Yfinal (=Y by default) was used to determine the final value of M based on the last iterated value of Ω)

### Locus-by-locus analysis for detecting differentially methylated CpG loci

We implemented a locus-by-locus analysis to identify differentially methylated CpG sites between samples obtained from control subjects without breast cancer diagnosis and those from healthy and diseased breasts before or after the cancer development using the R package *limma* [33]. Five groups were compared: 1) Controls with no breast cancer history, 2) Contralateral Prior Diagnosis (sample from healthy breast of a woman previously diagnosed breast cancer), 3) Ipsilateral Prior Diagnosis (sample from affected breast of a woman previously diagnosed breast cancer) 4) Contralateral New Diagnosis (sample from healthy breast of a woman with incident breast cancer), and 5) Ipsilateral New Diagnosis (sample from affected breast of a woman with incident breast cancer). Briefly, linear mixed effects models were fit to each CpG site separately, with the CpG β-value as the response against the five groups. A random effect for subject was included to control for within subject correlation in subjects with bilateral samples (30 subjects). The models were adjusted for time from delivery (in months), maternal age (in years), RefFreeCellMix proportion estimates (5 putative cell types), and the microarray Slide to control residual batch confounding. *P*-values were adjusted for multiple comparisons by computing the Benjamini–Hochberg *q*-values [34], and we defined loci with *q*-value < 0.05 to be statistically significant. We focus on CpGs identified as differentially methylated in both prospectively diagnosed groups (ipsilateral and contralateral), and report individual group results in supplemental material. All analyses were carried out using the R statistical package, version 3.5.0 (Vienna, Austria; www.r-project.org/) [35].

### Repetitive elements prediction and analysis

We use the package REMP, Repetitive Element Methylation Prediction [36], to estimate the DNA methylation levels on both *LINE-1* and *Alu* transposons using the information from the DNA methylation microarray. This random forest approach covers 37 *Alu* subfamilies and 115 *LINE-1* subfamilies. We computed the average *Alu* and *LINE-1* methylation levels for each sample, and tested the association with prospectively diagnosed breast cancer, excluding the three samples from subjects with a prior diagnosis of breast cancer. *P*-values were computed using the Kenward-Roger approach.

### Enrichment analyses

The probes that were differentially methylated were tested for pathway and gene set enrichment using missMethyl [37] and the MSigDB v.6.2 curated database [38]. A minimum of two genes were required for further exploring the specific pathway. We also tested for over- or underrepresentation of differentially methylated CpGs identified in the locus-by-locus analysis in 1) enhancer regions and 2) CpG island regions. Loci with a *q*-value < 0.05 were considered to be statistically significant. Odds ratios, 95% confidence intervals, and *P*-values were computed with the Cochran-Mantel-Haenszel test and were adjusted for probe type.

### Predicted methylation age and stem cell divisions

We used Horvath’s DNA methylation age estimation algorithm [39] to calculate predicted methylation age (*mAge*) using the agep function from wateRmelon [40]. Using those estimates, age acceleration was defined as: *Age acceleration = mAge - Age*. We tested for differences in age acceleration between control subjects and subjects with breast cancer using a linear mixed effects model. *P*-values were calculated using the Kenward-Roger approach. Additionally, stem cell divisions were estimated using the epiTOC method [41], but only 334 of 385 CpGs were available to calculate estimates. epiTOC estimates were compared between cases and controls using unadjusted linear mixed effect models analogously to the age acceleration models.

## RESULTS

Genome-scale DNA methylation was measured in breast milk samples from 87 subjects using the Illumina HumanMethylation450 beadchip. Subject demographic and sample details are provided in **Table 1**. 64 (73%) samples were from cancer-free subjects and 23 were from subjects who had a breast cancer diagnosis of which 20 (87%) were collected prior to diagnosis. Milk samples from subjects with any breast cancer diagnosis were classified according to whether the cancer was in the ipsilateral or contralateral breast. Overall, about 70% of samples from subjects with subsequent breast cancer were collected from the ipsilateral breast (n=14) and 30% were from the contralateral breast (n=6).

**Table 1.** Subject characteristics

| Variable | N (%) or Mean [Range] |  | P |
| --- | --- | --- | --- |
|  | Controls (n = 64) | Breast Cancer (n = 23) |  |
| Age (years) | 33.2 [23 - 44] | 36.3 [29 - 45] | 0.01 |
| BMI | 26.5 [18.2 - 43.6] | 25.2 [18.4 - 38.7] | 0.40 |
| BMI category |  |  | 0.20 |
| Normal/Underweight | 28 (43.8) | 8 (34.8) |  |
| Overweight/Obesity | 27 (42.2) | 13 (56.5) |  |
| Missing | 9 (14.1) | 2 (8.7) |  |
| Breast biopsy |  |  | <0.001 |
| No | 50 (78.1) | 0 (0.0) |  |
| Yes | 14 (21.9) | 23 (100.0) |  |
| Time since delivery (months) | 2.2 [0 - 10] | 10.8 [0.2 - 20] | <0.001 |
| Parity | 2 [1 - 5] | 2 [1 - 4] | <0.001 |
| Milk sample |  |  | N/A |
| Ipsilateral | N/A | 16 (69.6) |  |
| Contralateral | N/A | 7 (30.4) |  |
| Milk collection |  |  | N/A |
| Pre-diagnostic | N/A | 20 (87.0) |  |
| Post diagnosis | N/A | 3 (13.0) |  |

We used a reference-free cell type estimation approach to identify the number of putative cell types and the proportions of each cell type in each breast milk sample. The reference-free method identified five putative cell types in human milk. In unadjusted models, we observed differences in cell type proportions between breast milk samples from women who did not developed breast cancer (henceforth named as “controls”) compared with those diagnosed with breast cancer for three of the five putative cell types. The proportions of cell types 2 and 3 were higher in subjects with a prospective diagnosis of breast cancer than controls (*P*=5.2E-06 and 7.1E-04), and the proportion of cell type 4 was lower in milk from subjects with breast cancer compared to controls (*P*=1.2E-05) (**Figure 1**). In these models, differential abundance of putative cell types in controls versus cases was similar irrespective of whether the samples were from the ipsilateral or contralateral breast, or whether the breast cancer diagnosis occurred prior or subsequent to breast milk sample collection (see figure **Additional File 1**). After adjusting for maternal age (years), time since delivery (months), and BeadArray slide number, cell type proportions were no longer associated with breast cancer diagnosis.

**Figure 1.**
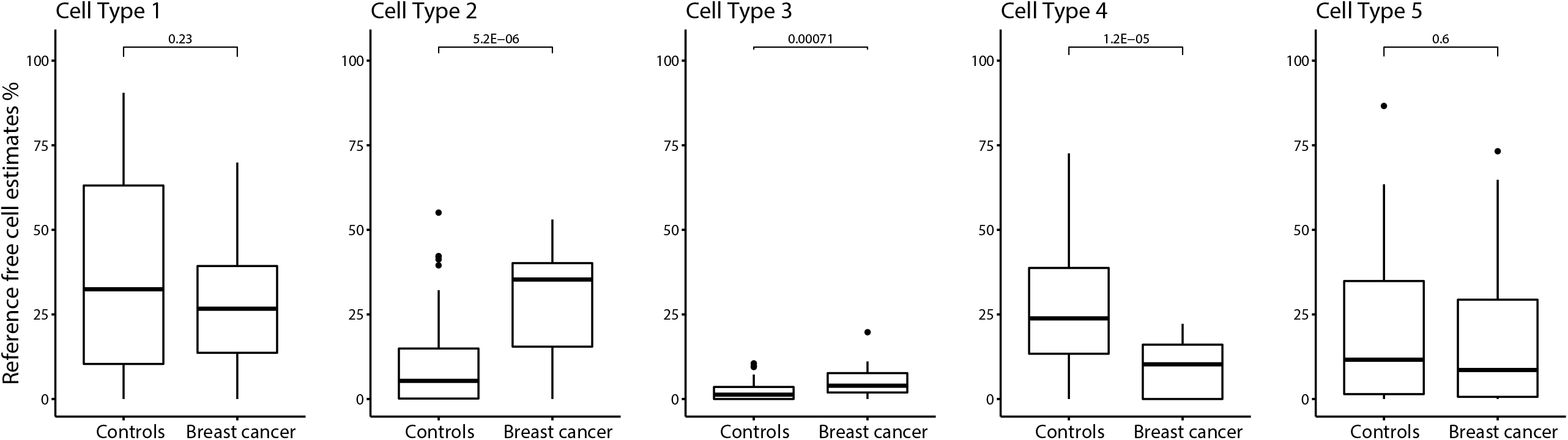
Percentage of reference-free cell estimates in subjects with and without breast cancer

DNA methylation was compared using linear mixed effect models adjusted for time since delivery in months, maternal age in years, estimated cell type proportions, and array chip with subject as a random effect. We identified 57 significantly differentially methylated CpG sites associated with milk from the ipsilateral breast after correction for multiple comparisons (*q-value* < 0.05). Among these 57 CpGs, one CpG in an island region and associated with both the *LRRC61* and *ACTR3C* genes was significantly hypermethylated in breast milk from subjects who were later diagnosed with breast cancer (**Figure 2**). The remaining 56 CpG sites were significantly hypomethylated in prospectively collected breast milk from the ipsilateral breast of subjects who developed cancer compared with controls (**Figure 2**). The most statistically significantly hypomethylated CpG site related to breast cancer diagnosis was located in the island region of the *CLCC1* gene. Additional genes with hypomethylated loci included *TMSB10, ZNF584, MAP10* (previously *KIAA1383), TRIM27*, and *SEPTIN7* (previously *SEPT7*). A total of 32 of these CpGs also were hypomethylated in prospectively collected milk from women who developed cancer in the contralateral breast compared to controls (**Table 2**). The full set of the EWAS results are available as **Additional Files 2 and 3**.

**Table 2.**
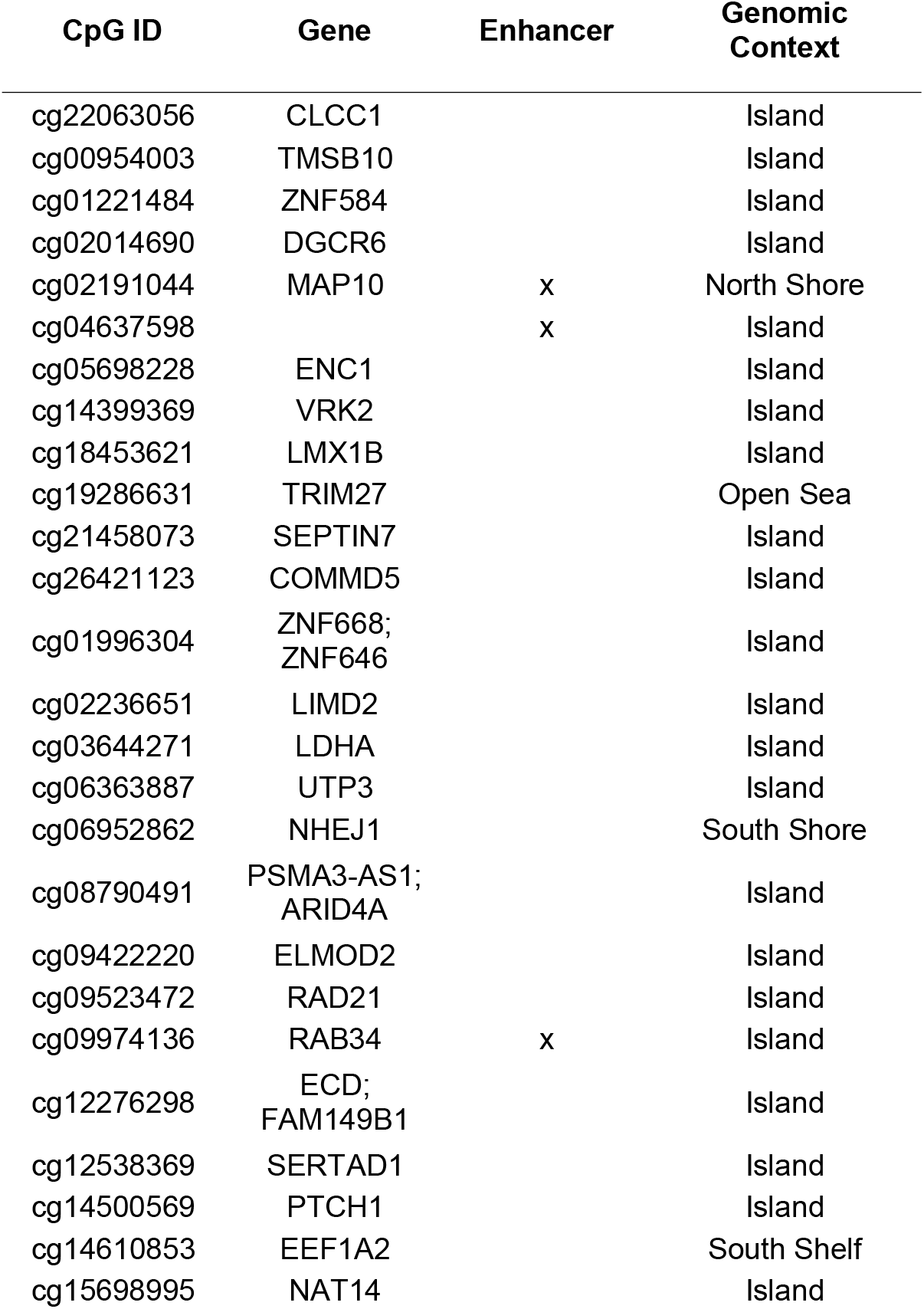
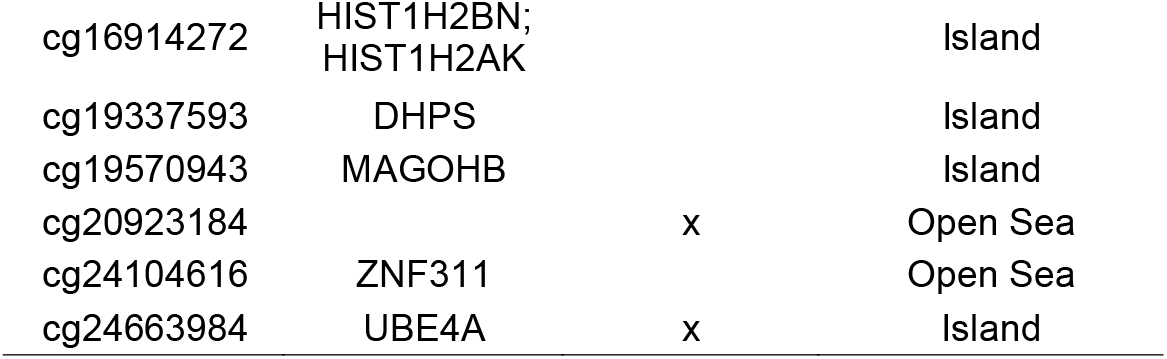
CpG loci that are hypomethylated in breast cancer

**Figure 2.**
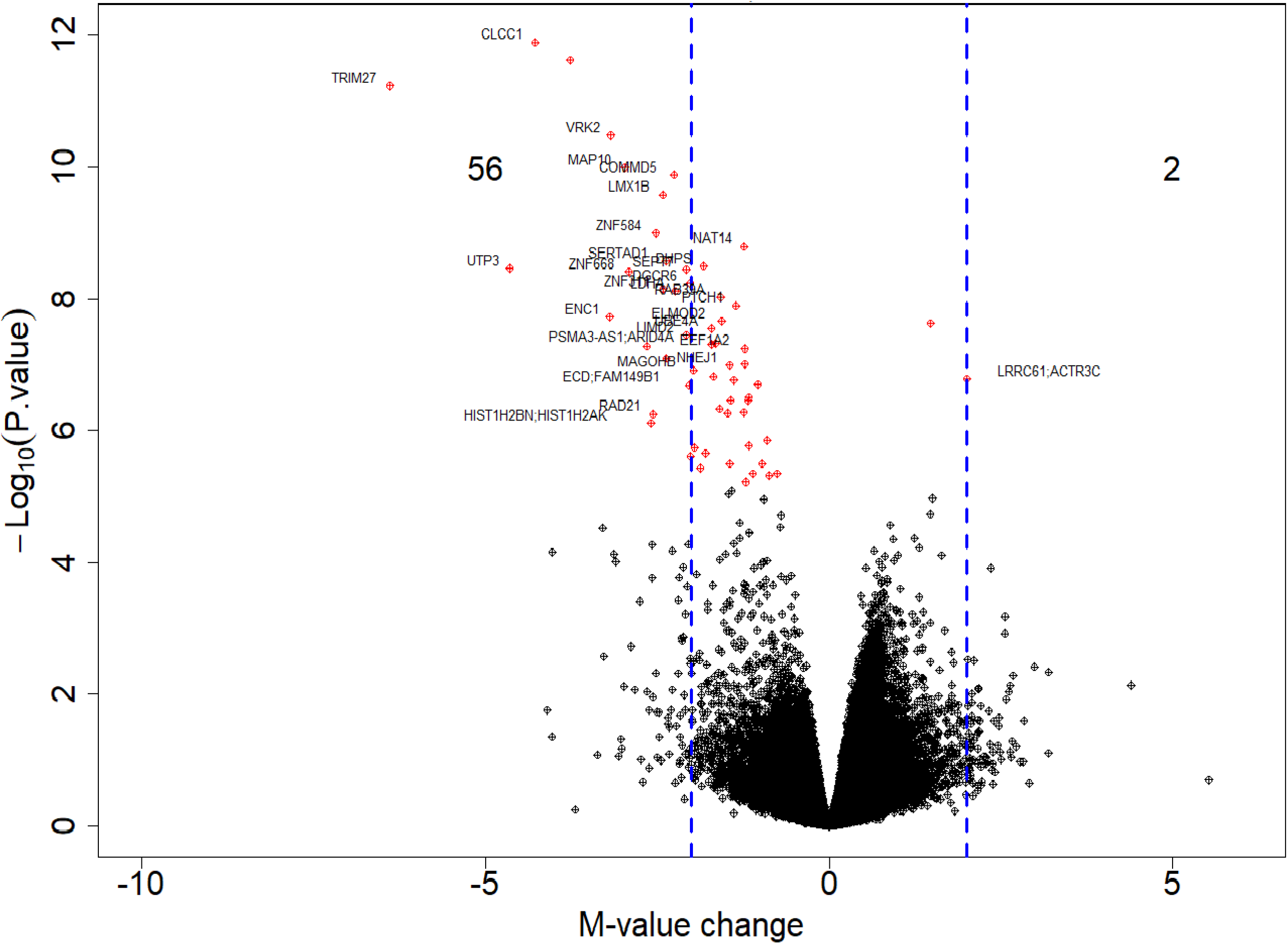
Volcano plots differentially methylated sites in milk from the ipsilateral breast in prospectively diagnosed cancer patients Note: In red those CpGs that were differentially methylated (q-value<0.05), 56 hypomethylated and 1 hypermethylated. Gene names were added to those CpG sites that overlapped with CpGs differentially methylated in the contralateral breast.

We accessed TCGA breast tumor data using cBioportal to determine whether genes we identified as having hypomethylated CpGs related to breast cancer were associated with gene regulation. We found negative correlations between DNA methylation with mRNA expression z-scores (RNA seq) for many of these genes including *ZNF584* (*P*=2.41E-17), *MAP10* (*P*=1.61E-76), *TRIM27* (*P*=6.01E-14), *LIMD2* (*P*=1.14E-59), and *LDHA* (*P*=6.06E-06). In contrast, there was little to no correlation between DNA methylation and expression of *CLCC1* (Spearman ρ=-0.03, *P*=0.5), *TMSB10* (ρ = −0.08, P=0.07) and *SEPTIN7* (ρ =-0.05, *P*=0.2), see **Additional File 4**. The range of DNA methylation level observed for each CpG tested in the TCGA tumors was comparable to that observed in our samples.

Given the preponderance of CpG-specific breast milk DNA hypomethylation associated with breast cancer, and that repeat element hypomethylation is well established in cancer, we further assessed repetitive element methylation. To do so, we inferred Alu (37 subfamilies) and LINE-1 (115 subfamilies) DNA methylation using array data and the repetitive element methylation prediction (REMP), as detailed in the methods section. None of the individual repetitive elements reached statistical significance after multiple comparison correction. The nominally significant are summarized in **Additional File 5, Table S5**. Mean Alu subfamily methylation was significantly lower in breast cancer cases compared to controls (β = −0.21, *p*-value = 2.9E-4), and mean LINE-1 subfamily methylation was also lower in cases than controls (β = −0.073, *p*-value = 0.10) (**Figure 3**).

**Figure 3.**
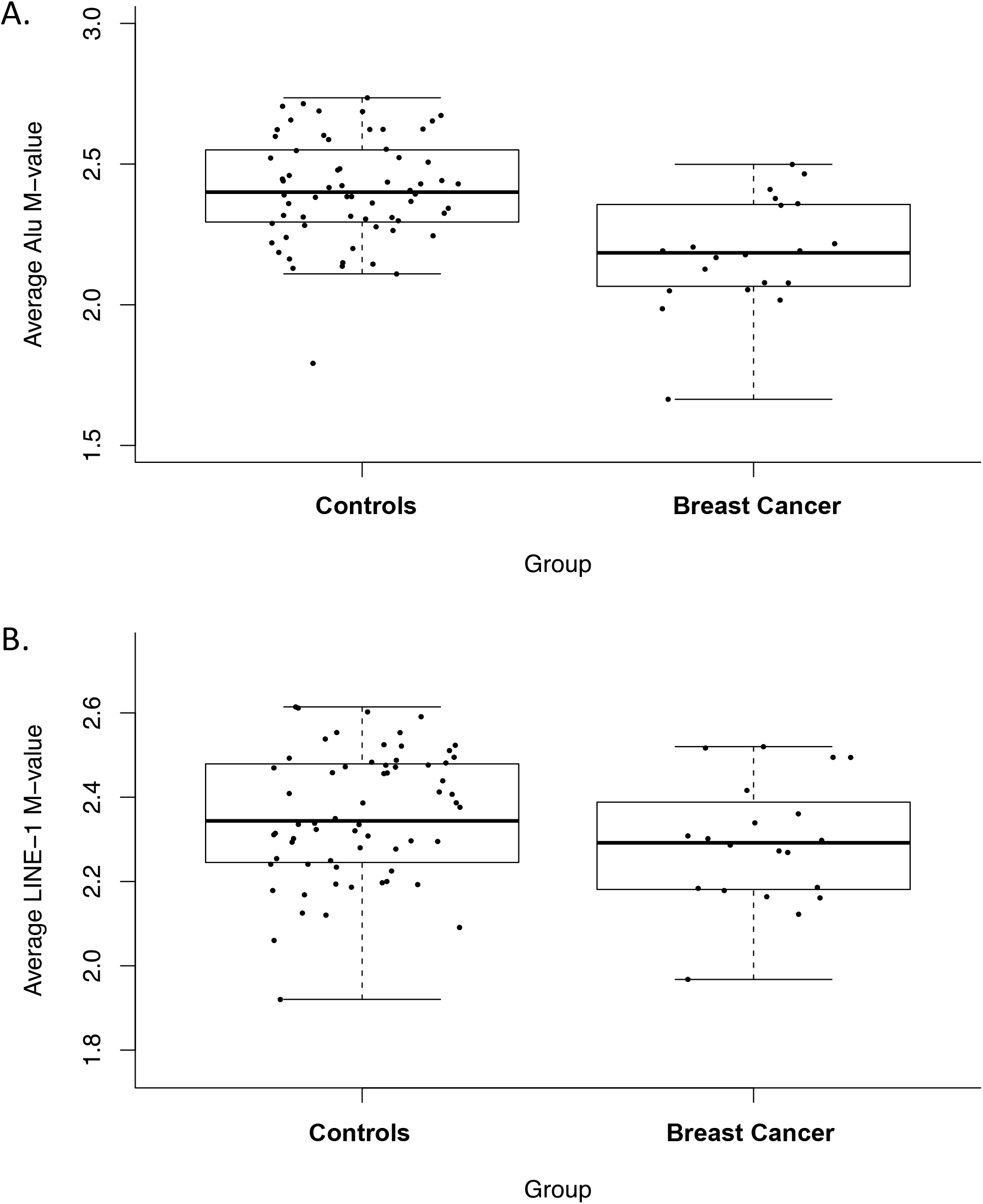
Differences in repetitive element CpG methylation by breast cancer status

To evaluate the location in the genome where breast cancer-related DNA methylation alterations in breast milk were occurring we performed enrichment analyses for both genomic context and gene sets. Differentially methylated CpGs (*q-value*<0.05) associated with a subsequent diagnosis of breast cancer were enriched for CpG island regions in milk from both the ipsilateral and contralateral breast (**Table 3**). Among CpGs whose methylation was significantly related with cancer diagnosis we also tested for enrichment of gene sets using the molecular signatures database (MSigDB) v. 6.2, and identified 7 gene sets enriched for the 32 CpG sites that were differentially methylated in both ipsilateral and contralateral samples. The top two pathways are related to highly conserved motif clusters matching transcription factor binding sites [42]. Three pathways are related to upregulation of genes in CD8(+) T lymphocytes, T regulatory cells and dendritic cells. Finally, two gene sets are associated to tumor invasion [43] and granulocyte differentiation in acute promyelocytic leukemia [44], see **Additional File 5, Table S6**.

**Table 3.** Enrichment for genomic context in CpGs with q < 0.05

| Breast Cancer Group<sup>2</sup> | Island Regions |  | Enhancer Regions |  |
| --- | --- | --- | --- | --- |
| | OR (95% CI) | $P^1$ | OR (95% CI) | $P^1$ |
| Ipsilateral | 3.48 (1.75, 7.45) | 9.3E-05 | 1.05 (0.45, 2.18) | 8.5E-01 |
| Contralateral | 4.28 (1.64, 13.30) | 8.6E-04 | 1.01 (0.30, 2.67) | 1.0E+00 |

<sup>1</sup> $P$ determined using the Cochran-Mantel-Haenszel test
<sup>2</sup>Reference level is controls with no breast cancer history

In univariate linear mixed effect analyses we also tested for DNA methylation age acceleration and elevated epigenetic mitotic clock tick rate (epiTOC) in association with breast cancer status. The epiTOC estimates were significantly higher amongst breast cancer subjects (β = 0.013, *p*-value = 3.2E-04, **Figure 4a**). A marginally significant increase in age acceleration subjects with breast cancer compared to controls was also observed (β = 2.7, *p*-value = 0.071, **Figure 4b**). After adjusting for covariates, neither epiTOC estimates nor methylation age were related to breast cancer status. In the adjusted analyses, putative cell type proportions were related to epiTOC estimates and age acceleration.

**Figure 4.**
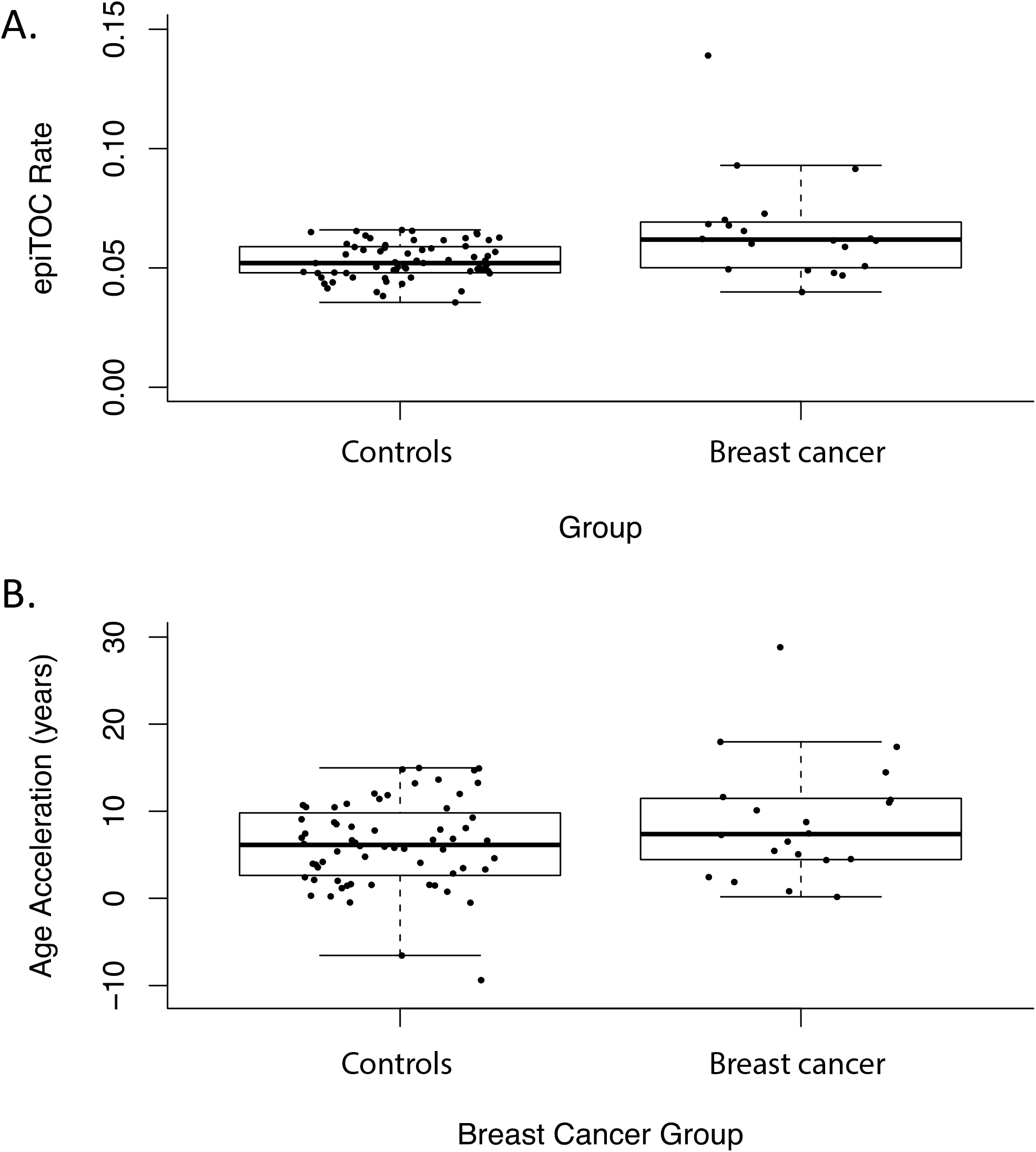
Measures of age inferred from methylation values

## DISCUSSION

We identified significant differences in DNA methylation after controlling for cell type and other confounders in subjects with a subsequent diagnosis of breast cancer compared with controls. In the subjects who were diagnosed with breast cancer after the milk collection, nearly all of the significantly differentially methylated CpGs were hypomethylated. Several of the genes whose CpG sites were differentially methylated in prospectively diagnosed cases have previously been associated with breast cancer. For example, *TMSB10* is overexpressed in breast cancer cells, has elevated protein expression in serum of breast cancer patients, and is elevated with increasing breast cancer stage and distant metastasis [45]. Linking a systemic marker of breast cancer risk to our tissue-specific approach, promoter CpG island hypomethylation of *ZNF584* was associated with a breast cancer diagnosis both here and in peripheral blood DNA from breast cancer patients [46]. Further, using TCGA breast tumor data, we showed the functional relationship of *ZNF584* DNA methylation with gene expression. We also observed hypomethylation at CpGs in *SEPTIN7, TRIM27, LIMD2*, and *LDHA*, which have been associated with breast cancer metastasis, invasion, and proliferation, [47–50]. Apart from *SEPTIN7*, all these genes showed negative correlation between gene expression and DNA methylation in TCGA breast cancer samples, again demonstrating functional consequences of altered DNA methylation to gene regulation. These results support our hypothesis that epigenetic alterations in human milk have utility for noninvasive molecular assessment of breast cancer risk.

Amongst subjects with incident breast cancer, the group of hypomethylated CpGs found to be significantly differentially methylated in milk samples from both contralateral and ipsilateral breast compared to those from controls were enriched for CpG island regions. Methylation at CpG island regions can reduce gene expression in associated genes [51]. Since the majority of differentially methylated CpGs were hypomethylated, this may correspond to increased expression of genes with promoters in these regions, and consistent with our observations of local and potentially systemic effects, our pathway enrichment analyses identified both proto-oncogene signatures and immune dysregulation signatures. One pathway with strong enrichment is associated with a motif for the *ELK-1* a regulator of the *c-Fos* protooncogene which has been linked to growth suppression in breast cancer cells [52]. The second pathway includes CpGs related to a motif for *SP-1*, a part of the Kruppel-like family that also has been associated as a prognostic factor in breast cancer [53]. Three more pathways pointed to genes upregulated in CD8(+) T lymphocytes, activated T-regulatory cells, and dendritic cells, cornerstones of tumor immune response in breast cancer murine models [54]. The remaining two pathways were related to tumor invasion and granulocyte differentiation.

We also observed differences in measures of methylation age including the epiTOC estimator and Horvath’s methylation age between breast cancer subjects and control subjects, but these associations were not robust to adjustment for potential confounders. Notably, in our study population the subjects with a cancer diagnosis were slightly older than control subjects. Putative cell type proportions, however, were related to all measures of methylation age. Although CpG loci utilized in each algorithm are not supposed to be dependent on cell type, there have been consistent trends of accelerated age in breast tissues when using the Horvath methylation age approach. Accelerated biologic age inferred using DNA methylation has recently been associated with breast cancer risk in a very large prospective study using peripheral blood [55]. However, to date, unlike peripheral blood, there are no DNA methylation clocks for inference of biologic age that are calibrated to biospecimens from the breast. In the future, larger breast-tissue-specific studies are needed to advance our understanding and opportunity to leverage biologic age estimates for breast cancer risk assessment and primary prevention.

This study has several strengths and limitations. Strengths of our study include the use of prospectively collected specimens, tissue-specific measures of DNA methylation, and two independent cohorts. Although some subjects were potentially had clinically occult disease when providing a milk specimen, others were not diagnosed until years later. One limitation of this study is sample size, though investigating genome-scale DNA methylation measures in breast milk is novel. One potential limitation is that we pooled controls from two different cohorts processed in different technical batches. Although we controlled for technical differences in our models and used a conservative approach that adjusted for cell estimates which also captures technical differences, we cannot completely exclude some residual technical noise between cohorts affecting our results.

We identified early DNA methylation alterations in breast milk associated with subsequent breast cancer occurrence. These loci were either in genes expressed in breast cancers, related to breast cancer progression, or found in peripheral blood samples women with breast cancer. Importantly, because we identified both overlapping results with work that used peripheral blood as a surrogate biospecimen and results distinct to breast milk we expect that our tissue-specific approach has high potential for follow up work. We expect that future investigations of DNA methylation changes present in cells from breast milk from disease-free women will have value for risk assessment and primary prevention of breast cancer, perhaps with specific strength in application to premenopausal disease. However, larger studies are needed to validate our findings and to further establish the utility of breast milk as a biospecimen for understanding the molecular basis of disease risk and prospective risk assessment.

## Conclusions

We assessed genome wide DNA methylation in breast milk from subjects with and without breast cancer, specific loci were hypomethylated in breast cancer subjects compared to control subjects. These differentially methylated regions were more likely to occur in island regions of the genome. Our results suggest that breast milk has utility for prospective assessment of breast cancer risk.

## Data Availability

The datasets generated and analyzed during the current study are available in the GEO (https://www.ncbi.nlm.nih.gov/geo/) under the accession number GSE133918.

## List of abbreviations

EWAS: Epigenome-wide association studies
epiTOC: epigenetic mitotic clock tick rate
MSigDB: molecular signatures database
UMass: University of Massachusetts Amherst
NHBCS: New Hampshire Birth Cohort study
*mAge*: methylation age

## Declarations

### Ethics approval and consent to participate

All study participants provided written informed consent prior to the study according to the guidelines of Institutional Review Board of the University of Massachusetts Amherst and the Committee for the Protection of Human Subjects at Dartmouth.

### Consent for publication

Not applicable

## Authors’ contributions

LAS and SNL elaborated the analysis plan, analyzed the data and wrote the first draft of the manuscript, EPB, ECP, DLA, MRK provided technical and methodological feedback to the original analysis and final version, KFA and BCC generated the original idea and codirected the analyses. All authors approved the final version of this manuscript.

## Acknowledgements

Not applicable.

## Additional files

**Additional File 1: Figure S1**. Percentages of reference-free cell estimates by breast cancer group

**Additional File 2: Table S1:** EWAS results contralateral breast milk new cancer diagnosis after milk donation. **Table S2:** EWAS results ipsilateral breast milk new breast cancer diagnosis after milk donation. **Table S3:** EWAS results contralateral breast milk previous breast cancer. **Table S4:** EWAS results ipsilateral breast milk previous breast cancer.

**Additional File 3: Figure S2**. Volcano plots differentially methylated sites in breast milk according to breast source cancer status

**Additional File 4: Figure S3**. Correlation between gene expression (mRNA) and DNA methylation in breast tumor samples from TCGA

**Additional File 5: Table S5**. LINE-1 element methylation in realtion to breast cancer diagnosis (P-value < 0.01). **Table S6**. Molecular Signatures Database (MSigDB) pathways enriched using missMethyl

